# Safety and efficacy of the ChAdOx1 nCoV-19 (AZD1222) Covid-19 vaccine against the B.1.351 variant in South Africa

**DOI:** 10.1101/2021.02.10.21251247

**Authors:** Shabir A. Madhi, Vicky Baillie, Clare L. Cutland, Merryn Voysey, Anthonet L. Koen, Lee Fairlie, Sherman D. Padayachee, Keertan Dheda, Shaun L. Barnabas, Qasim Ebrahim Bhorat, Carmen Briner, Gaurav Kwatra, NGS-SA, Wits-VIDA COVID team, Khatija Ahmed, Parvinder Aley, Sutika Bhikha, Jinal N. Bhiman, As’ad Ebrahim Bhorat, Jeanine du Plessis, Aliasgar Esmail, Marisa Groenewald, Elizea Horne, Shi-Hsia Hwa, Aylin Jose, Teresa Lambe, Matt Laubscher, Mookho Malahleha, Masebole Masenya, Mduduzi Masilela, Shakeel McKenzie, Kgaogelo Molapo, Andrew Moultrie, Suzette Oelofse, Faeezah Patel, Sureshnee Pillay, Sarah Rhead, Hylton Rodel, Lindie Rossouw, Carol Taoushanis, Houriiyah Tegally, Asha Thombrayil, Samuel van Eck, Constantinos Kurt Wibmer, Nicholas M. Durham, Elizabeth J Kelly, Tonya L Villafana, Sarah Gilbert, Andrew J Pollard, Tulio de Oliveira, Penny L. Moore, Alex Sigal, Alane Izu

**Affiliations:** South African Medical Research Council Vaccines and Infectious Diseases Analytics Research Unit, Faculty of Health Sciences, University of the Witwatersrand, Johannesburg, South Africa; Department of Science and Innovation/National Research Foundation South African Research Chair Initiative in Vaccine Preventable Diseases Unit, University of the Witwatersrand, Johannesburg, South Africa; African Leadership in Vaccinology Expertise, Faculty of Health Sciences, University of the Witwatersrand, Johannesburg, South Africa; Oxford Vaccine Group, Department of Paediatrics, University of Oxford, UK; Wits Reproductive Health and HIV Institute, Faculty of Health Sciences, University of the Witwatersrand, Johannesburg, South Africa; Setshaba Research Centre, Tshwane, South Africa; Division of Pulmonology, Groote Schuur Hospital and the University of Cape Town, South Africa Faculty of Infectious and Tropical Diseases, Department of Immunology and Infection, London School of Hygiene and Tropical Medicine, London, UK; Family Centre for Research with Ubuntu, Department of Paediatrics, University of Stellenbosch, Cape Town, South Africa; Soweto Clinical Trials Centre, Soweto, South Africa; Perinatal HIV Research Unit, Faculty of Health Sciences, University of the Witwatersrand, Johannesburg, South Africa; Network for Genomic Surveillance in South Africa (NGS-SA) group; Wits VIDA COVID vaccine trial group; Astra Zeneca Biopharmaceuticals, Cambridge, UK; Jenner Institute, Nuffield Department of Medicine, University of Oxford, UK; National Institute for Communicable Diseases (NICD) of the National Health Laboratory Service (NHLS), Johannesburg, South Africa; Antibody Immunity Research Unit, School of Pathology, Faculty of Health Sciences, University of the Witwatersrand, Johannesburg, South Africa; Africa Health Research Institute, Durban, South Africa; Max Planck Institute for Infection Biology, Berlin, Germany; Division of Infection and Immunity, University College London, London, UK; KwaZulu-Natal Research and Innovation Sequencing Platform (KRISP), University of KwaZulu-Natal, Durban South Africa

## Abstract

**Background:** Assessing safety and efficacy of Covid-19 vaccines in different populations is essential, as is investigation of efficacy against emerging SARS-CoV-2 variants of concern including the B.1.351 (501Y.V2) variant first identified in South Africa.

**Methods:** We conducted a randomized multicentre, double blinded controlled trial on safety and efficacy of ChAdOx1-nCoV19 in HIV-uninfected people in South Africa. Participants age 18 to <65 years randomized (1:1) to two doses of vaccine containing 5×10^10^ viral particles or placebo (0.9%NaCl) 21-35 days apart. Post 2^nd^-dose serum samples (n=25) were tested by pseudotyped (PSVNA) and live virus (LVNA) neutralization assays against the D614G and B.1.351 variants. Primary endpoints were safety and vaccine efficacy (VE) >14 days following second dose against laboratory confirmed symptomatic Covid-19.

**Results:** 2026 HIV-uninfected adults were enrolled between June 24^th^ and Nov 9^th^, 2020; 1010 and 1011 received at least one dose of placebo or vaccine, respectively. Median age was 31 years. The B.1.351 variant showed increased resistance to vaccinee sera using the PSVNA and LVNA. In the primary endpoint analysis, 23/717 (3.2%) placebo and 19/750 (2.5%) vaccine recipients developed mild-moderate Covid-19; VE 21.9% (95%Confidence Interval: −49.9; 59.8). Of the primary endpoint cases, 39/42 (92.9%) were the B.1.351 variant; against which VE was 10.4% (95%CI: −76.8; 54.8) analyzed as a secondary objective. The incidence of serious adverse events was balanced between the vaccine and placebo groups.

**Conclusions:** A two-dose regimen of ChAdOx1-nCoV19 did not show protection against mild-moderate Covid-19 due to B.1.351 variant, however, VE against severe Covid-19 is undetermined.

(Funded by The Bill & Melinda Gates Foundation and South African Medical Research Council; ClinicalTrails.gov number, NCT04444674).

## Background

There has been unprecedented speed in developing vaccines for Covid-19, which is caused by the severe acute respiratory syndrome coronavirus-2 (SARS-CoV-2), since being declared a pandemic by World Health Organization (WHO) on March 11, 2020.^1-4^ ChAdOx1-nCoV19, a replication-deficient chimpanzee adenoviral vector containing the SARS-CoV-2 structural surface glycoprotein antigen, is one of six COVID-19 vaccines based on different platforms that have been authorized for emergency use,^5-11^ with efficacy results of two further vaccines having recently been reported.^12,13^ Thus far, excluding the South African components of the Novavax^12^ and Jansen Covid-19 vaccine trials^13^ and the inactivated whole virus vaccines,^10^ preclinical and most clinical studies assessed protection against the prototype SARS-CoV-2 sequence (B.1) or the D614G variant.^5,6,8,9^

Recently, the SARS-CoV-2 spike genome has accumulated mutations, including within the receptor binding domain (RBD) and N-terminal domains (NTD)^.14,15^ These domains are major targets of the antibody response. The RBD mutations include the N501Y mutation which is associated with increased affinity for the angiotensin converting enzme-2 (ACE2).^16^ In contrast, E484K and K417N RBD mutations and mutations in the NTD have been associated with neutralizing antibody escape.^17^ The B.1.1.7 lineage, first identified in the United Kingdom (UK), includes the N501Y mutation which has been associated with 53% increased transmissibly.^18^ Neutralizing antibody activity elicited by infection or mRNA vaccines against the B.1.1.7 variant are largely unaffected.^19^ The B.1.1.7 variant has, however, now evolved to include the E484K mutation in the UK^.20^

The B.1.351 (N501Y.V2) lineage first identified in South Africa contains the three RBD mutations and five additional NTD mutations^.14,15^ The sensitivity of B.1.351 to neutralizing antibodies from convalescent donors infected with the prototype lineage virus using a spike-pseudotyped neutralization (PSVNA) assay demonstrated 48% of sera were unable to neutralize B.1.351, with the rest showing 3 to 86 fold reduction in neutralization titres.^21^ This was corroborated using a live virus neutralization assay (LVNA), with reduction in antibody activity ranging from 6-fold to complete knockout for the B.1.351 variant.^14^ Another independent lineage of SARS-CoV-2 (P.1) also containing the E484K, K417N and some B.1.351 NTD mutations has been identified in Brazil^.22,23^

An interim pooled analysis of ChAdOx1-nCoV19 efficacy in the UK and Brazil, prior to evolution of the B.1.351 and P.1 variants, reported overall vaccine efficacy (VE) of 70.4% (95.8% Confidence interval [CI] 54.8; 80.6).^8^ Recent analysis of ChAdOx1-nCoV19 VE against the B.1.1.7 (N501Y.V1) in the UK was 74.6% (95%CI 41.6-88.9).^24^

Here, we report on a multi-centred South African phase Ib/II trial evaluating the safety, immunogenicity and VE of ChAdOx1-nCoV19 in preventing symptomatic COVID-19. This interim analysis is limited to addressing co-primary safety analysis; and primary and key secondary VE objectives including efficacy specifically against B.1.351 variant. Furthermore, we report on immunogenicity of ChAdOx1-nCoV19, and post-hoc PSVNA and LVNA investigation of the sensitivity of the original D614G virus and the B.1.351 variant to vaccine-elicited antibodies.

## METHODS

### Trial objectives, participants and oversight

In this randomized, double-blinded, placebo-controlled, multi-site trial conducted in South Africa, we assessed the safety and efficacy of two standard doses of ChAdOx1-nCoV19 administered 21-35 days apart, compared to saline (0.9% NaCl) placebo. Adults aged 18 to <65 years, with no or well-controlled chronic medical conditions, were eligible for participation. Key exclusion criteria included HIV positivity at screening (for the efficacy cohort), previous or current laboratory-confirmed Covid-19, history of anaphylaxis in relation to vaccination and morbid obesity (body mass index ≥40kg/m^2^). Detailed inclusion and exclusion criteria are found in Supplementary text 1.1. ChAdOx1-nCoV19 was developed at the University of Oxford, who sponsored the trial, with sources of study vaccine supply reported in Supplement text 1.2.

Trial data were available to all authors, who confirm its accuracy and completeness. An independent data and safety monitoring committee reviewed efficacy and unblinded safety data. A local trial safety physician reviewed all serious adverse events in real-time. The trial was monitored by an external clinical research organization, who ensured adherence to the protocol (appendix 1).

The trial was reviewed and approved by the South African Health Products Regulatory Authority (SAHPRA, ref 20200407), ethics committees of the University of the Witwatersrand (ref 200501), Cape Town (ref 350/2020), Stellenbosch (ref M20/06/009_COVID-19) and Oxford (ref 35-20) prior to trial initiation and was registered on Clinicaltrials.gov (NCT04444674) and the Pan African Clinical Trials registry (PACTR202006922165132). All participants were fully informed about trial procedures and possible risks and signed informed consent prior to enrolment.

### Trial procedures

A cohort of 70 HIV-uninfected individuals were enrolled first (Group-1) for intensive safety and immunogenicity monitoring, followed by enrolment of another 1956 HIV-uninfected individuals, all (excluding five who were not vaccinated) were included in safety analysis and 1467 for the primary efficacy analysis (Fig. 1). Details of study procedures are in the protocol (Appendix 1; pg 68-73). The study follow-up is ongoing.

Trial participants were randomized to receive either 0.33-0.5ml ChAdOx1-nCoV19 (lot dependent) or placebo intramuscularly into the deltoid muscle of the non-dominant arm on day of randomization and a booster 21-35 days later. Participants were observed for 30 minutes post vaccination for acute reactions. Study-injection preparation and vaccination were done by unblinded site staff who were not involved in any other study procedures. Trial participants and all other study staff remain blinded to treatment group.

### Safety

The safety analysis evaluated occurrence of solicited local and systemic reactogenicity within 7 days following vaccination, unsolicited adverse events for 28 days following vaccination, changes in baseline safety laboratory measures (Group-1 only) and serious adverse events. Further details of evaluating for safety and reactogenicity are detailed in Supplementary Text 1.3. Adverse event data reported up until January 15^th^, 2021 are included in this report.

### SARS-CoV-2 testing, whole genome sequencing and genome assembly

Testing for SARS-CoV-2 infection using a nucleic acid amplification test (NAAT) included sampling at routine scheduled visits detailed in the protocol (Appendix 1; pages 68-73); as well as when participants presented with any symptoms suggestive of Covid-19. Participants were advised at time of randomization as to which clinical symptoms suggestive of Covid-19 should trigger them to present to be investigated for SARS-CoV-2 infection (Table S1). Also, short message service was sent every two weeks as a reminder to present for investigation if symptomatic. Details of NAAT, whole genome sequencing and phylogenetic analysis are described in Supplementary Text 1.4.

### Neutralization assays

SARS-CoV-2 serostatus at randomization was evaluated using an IgG assay to the nucleoprotein (N) as described.^8^ For antibody neutralization studies, PSVA (methods in Supplementary 1.5) to prototype virus was performed on serum samples obtained two weeks after the 2^nd^ dose of vaccine in 107 randomly selected ChAdOx1-nCoV19 recipients who were seronegative for IgG N-protein at enrolment. In addition, we present post-hoc data on the comparative pseudoneutralization for vaccine recipients enrolled in the Brazil (n=226) and UK (n=326) ChAdOx1-nCoV19 studies.

To assess neutralization activity of vaccine-elicited antibodies against B.1.351, 25 serum samples from Group-1 participants who tested seronegative at enrolment but showed varying neutralizing antibody titers at 14 days post 2^nd^ dose of injection were assayed using PSVNA and LVNA before unblinding of study arm allocation^.14,21^ The PSVNA tested neutralization activity against the original D614G spike, a RBD triple mutant (containing only K417N, E484K, N501Y) and the B.1.351 spike. Live virus neutralization assay was performed by a microneutralization focus forming assay in Vero E6 cells. Details of the PSVA and LVNA has been published and are briefly described online; Supplementary Text 1.5^.14,21^

### Efficacy objectives

Covid-19 cases were evaluated by at least two physicians who were independent of the study and masked to study-arm assignment, discordant assessments were evaluated by a third adjudicator. Grading of severity was based on a pre-specified scoring system; Supplementary Table S1 and S2.

The primary endpoint was efficacy against NAAT-confirmed symptomatic Covid-19 occurring more than 14 days after the 2^nd^ injection in participants who were seronegative at randomization. VE against B.1.351 lineage was a pre-specified secondary objective.

Other secondary efficacy objectives included VE against Covid-19 for the overall population (inclusive of seropositive participants at randomization), VE specific to baseline seropositive group, and VE against Covid-19 occurring >14 or >21 days after the first dose. Further details of secondary VE analyses are included in Appendix 1 (pg: 41-48). Furthermore, a post-hoc analysis was done for the overall and seronegative population, to evaluate VE occurring >14 days after the first injection with endpoint cases restricted until 31^st^ October 2020 as a proxy for non-B.1.351 variant Covid-19. The B.1.351 variant only started to be identified in the areas where the study sites (Johannesburg and Tshwane in Gauteng, and Cape Metro in Western Cape Province) were based from mid-November 2020 onward; Supplementary Figure S1^.15^

### Statistical analysis

Participants who received at least one dose of ChAdOx1 nCoV-19or placebo and returned diary cards completed until day 7 post first vaccination were included in the safety reactogenicity analysis. The occurrence of each solicited local- and systemic reactogenicity sign and symptom for 7 days following vaccination, adverse events, and SAEs up to January 15, 2021 are presented stratified by arm.

For the primary efficacy analyses, only per-protocol seronegative participants were included. VE was calculated as 1-the relative risk and 95% CI were calculated using the Clopper-Pearson exact method are reported. A sensitivity analysis was conducted which includes modified intention-to-treat, seronegative participants irrespective of whether they received the vaccine or placebo.

## RESULTS

### Participants

We screened 3,022 individuals across seven sites, of whom 2,026 HIV-uninfected individuals were enrolled from June 24^th^ through to November 9^th^, 2020. The initiation of enrolment coincided with the peak of the first Covid-19 wave in South Africa; Supplementary Figure S2. Overall, 1,010 participants received vaccine and 1,011 received placebo (Fig. 1). There were 1,467 (750 vaccinees and 717 placebo) Covid-19-naïve participants eligible for the primary VE analysis, reason for exclusion indicated in Figure 1.

The median age was Figure 131 years, 56.5% identified as male gender, and the racial distribution included 70.5% black-Africans, 12.8% whites and 14.9% identifying as “mixed” race. Nineteen percent of enrolees were obese (BMI≥30-40 kg/m^2^), 42.0% were smokers, 2.8% had underlying hypertension and 3.1% had chronic respiratory conditions. The median duration between doses was 28 days; and the median duration of follow-up from enrolment and from 14 days after the second dose of injection were 156 and 121 days, respectively (as of January 15^th^, 2021). Demographics of the baseline seronegative population was like the overall population; Table 1.

**Table 1:** Baseline demographic and other characteristic of overall study population contributing to the safety analysis, and population contributing to the primary efficacy endpoint analysis.

| Variable | Overall safety study population<sup>a</sup> |  |  | Seronegative efficacy study population<sup>b</sup> |  |  |
| --- | --- | --- | --- | --- | --- | --- |
|  | Total | Placebo | Vaccine | Total | Placebo | Vaccine |
| N enrolled | 2021 | 1010 | 1011 | 1467 | 717 | 750 |
| Male n (%) | 1142 (56.5) | 568 (56.2) | 574 (56.8) | 838 (57.1) | 397 (55.4) | 441 (58.8) |
| Median Age in years (IQR) | 30 (24-40) | 30 (24-39) | 31 (24-40) | 31 (24-41) | 31 (24-41) | 31 (24-41) |
| Age in years; n (%) |  |  |  |  |  |  |
| 18-<45 | 1695 (83.9) | 852 (84.4) | 843 (83.4) | 1206 (82.2) | 593 (82.7) | 613 (81.7) |
| 45-<60 | 283 (14) | 133 (13.2) | 150 (14.8) | 223 (15.2) | 102 (14.2) | 121 (16.1) |
| >=60 | 43 (2.1) | 25 (2.5) | 18 (1.8) | 38 (2.6) | 22 (3.1) | 16 (2.1) |
| BMI; n (%)<sup>c</sup> |  |  |  |  |  |  |
| 0-<18.5 | 151 (7.5) | 68 (6.7) | 83 (8.2) | 119 (8.1) | 50 (7) | 69 (9.2) |
| 18.5 - <25 | 1021 (50.6) | 521 (51.6) | 500 (49.6) | 752 (51.4) | 371 (51.7) | 381 (51.0) |
| 25 - <30 | 456 (22.6) | 221 (21.9) | 235 (23.3) | 330 (22.5) | 156 (21.8) | 174 (23.3) |
| >=30 | 390 (19.3) | 200 (19.8) | 190 (18.8) | 263 (18.0) | 140 (19.5) | 123 (16.5) |
| Smoker; n (%) | 849 (42) | 415 (41.1) | 434 (42.9) | 644 (43.9) | 304 (42.4) | 340 (45.3) |
| Alcohol; n (%) | 990 (49) | 501 (49.6) | 489 (48.4) | 729 (49.7) | 365 (50.9) | 364 (48.5) |
| Health worker; n (%) | 167 (8.3) | 88 (8.7) | 79 (7.8) | 144 (9.8) | 80 (11.2) | 64 (8.5) |
| Race; n (%) |  |  |  |  |  |  |
| Black African; n (%) | 1421 (70.5) | 708 (70.) | 713 (70.6) | 949 (64.9) | 453 (63.4) | 496 (66.2) |
| Mixed | 300 (14.9) | 149 (14.8) | 151 (15) | 251 (17.2) | 128 (17.9) | 123 (16.4) |
| White | 259 (12.8) | 132 (13.1) | 127 (12.6) | 231 (15.8) | 119 (16.7) | 112 (15.0) |
| Other | 37 (1.8) | 18 (1.8) | 19 (1.9) | 32 (2.2) | 14 (2.0) | 18 (24) |
| Hypertension; n (%) | 56 (2.8) | 25 (2.5) | 31 (3.1) | 42 (2.9) | 20 (2.8) | 22 (2.9) |
| Respiratory system;<br>n (%) | 62 (3.1) | 26 (2.6) | 35 (3.6) | 53 (3.6) | 22 (3.1) | 31 (4.1) |
| Diabetes; n (%) | 9 (27.3) | 5 (23.8) | 4 (36.4) | 5 (19.2) | 3 (17.6) | 2 (22.2) |
| Median days<br>between doses;<br>(IQR) | 28 (28-32) | 28 (28-32) | 28 (28-32) | 28 (28-32) | 28 (28-32) | 28 (28-32) |
| Median days of<br>follow-up from<br>randomization (IQR) | 156<br>(140-171) | 156<br>(140-171) | 156<br>(140-171) | 161<br>(143-172) | 160<br>(142-172) | 161<br>(143-174) |
| Median days of 2<sup>nd</sup><br>injection (IQR) | 121<br>(114-143) | 121<br>(114-142) | 121<br>(114-143) | 122<br>(114-143) | 122<br>(114-142) | 128<br>(114-143) |
| Per person days of<br>follow-up from<br>randomization | 290394 | 143962 | 146432 | 229129 | 111471 | 117658 |
| Per person days of<br>follow-up from 2<sup>nd</sup><br>injection | 228506 | 113063 | 115443 | 184595 | 89714 | 94881 |

<sup>a</sup>Inclusive of all participants who received at least one dose of vaccine or placebo and irrespective of baseline sero-status suggestive of past SARS-CoV-2 infection or NAAT positivity within 96 hours and on day of randomization. Excluding five participants whom were randomized to treatment arm and never received a placebo or vaccine
<sup>b</sup> Population included in vaccine efficacy analysis for the primary objective, who had a non-reactive NAAT within 96 hours and on day of randomization, and tested negative for SARS-CoV-2 N-protein IgG.
<sup>c</sup>BMI= Body Mass Index in kilograms per square meter.
IQR=Inter-quartile range

### Safety

Local and systemic reactogenicity data are presented in Supplementary Figures S3 and S4 and summarised in Supplementary Text 1.6. Adverse event and serious adverse event rates were similar amongst vaccine and placebo recipients; Supplementary Table S3 and S4. The only serious adverse event attributed to ChAdOx1-nCoV19 was a participant with temperature of >40°C after the 1^st^ dose which subsided within 24 hours, and in whom no reactogenicity was observed after the 2^nd^ dose. All other events were considered unrelated or unlikely to be related to injection received.

### Immunogenicity

#### Pseudotyped and live virus neutralization assays

Humoral response to ChAdOx1-nCoV19 induced strong neutralising antibodies at 28 days after the first dose, which rose further after a second dose. The responses in ChAdOx1-nCoV19 recipients in our study was similar those from the UK and Brazil studies (Fig. 2a).

**Figure 2:**
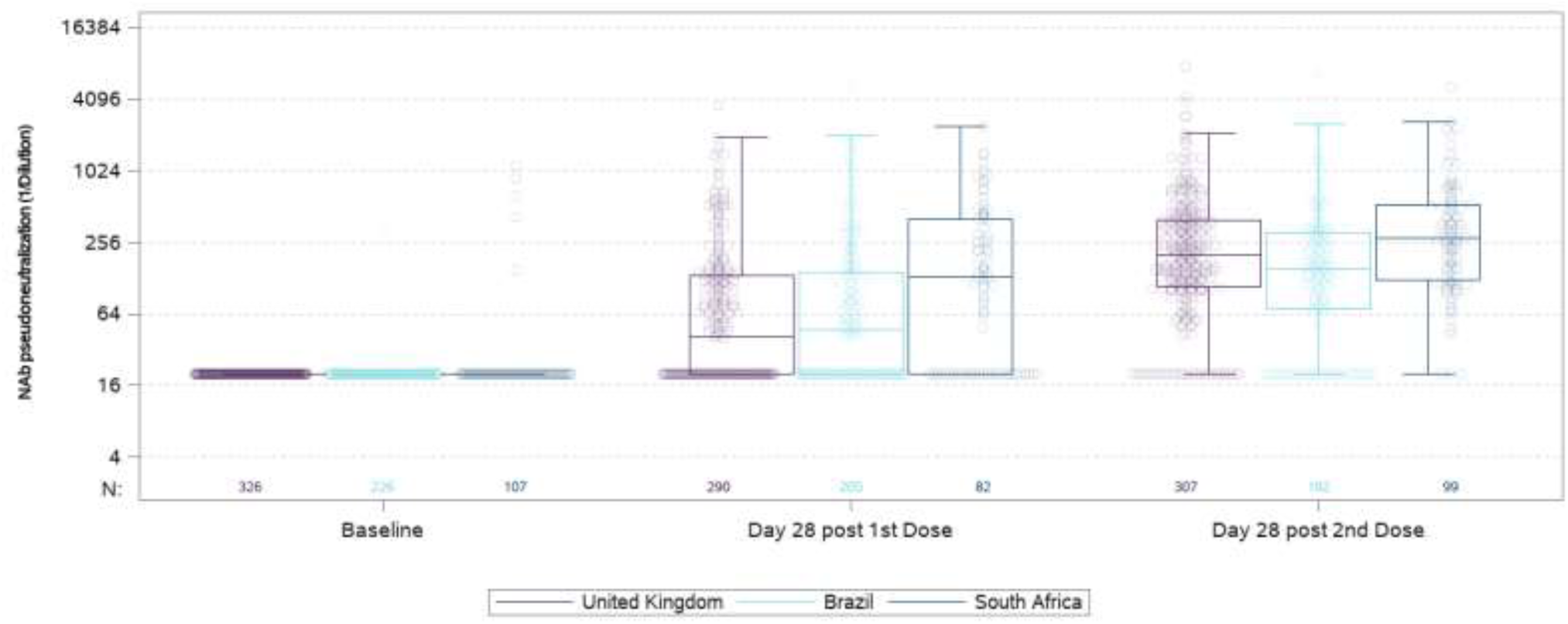
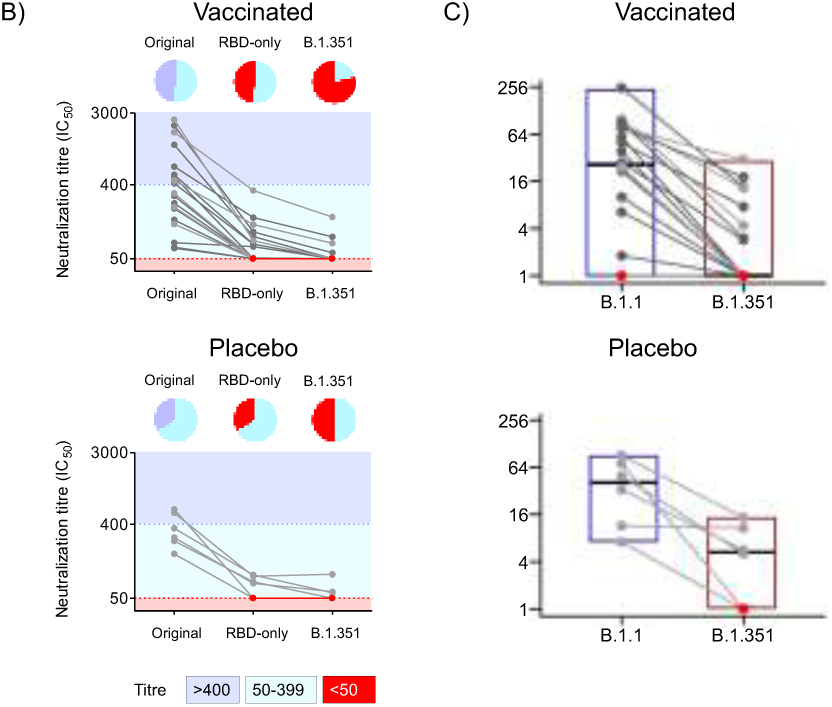
Pseuduoneutralization assay to original SARS-CoV-2 virus in ChAdOx1 nCoV-19 (AZD1222) recipients from the United Kingdom, Brazil and South Africa; Fig 2b: Pseduneutralization assay to original, triple recpetor-binding domain and B.1.351 variant; Fig 2c Live virus neutralization virus assay against original and B.1351 SARS-CoV-2. Fig 2a: Vaccine serum from baseline seronegative 18-64 year old AZD1222 vaccinees receiving two standard doses in the United Kingdom (n=326), Brazil (n=226), and South Africa (n=107) were evaluated in a validated pseudoneutralisation assay at baseline, 28 days after first dose and 28 days after second dose. Boxes show median (IQR). Note: The AZD1222-recipients included in the analysis were randomly seleceted partcipants from the efficay trial, who contributed to the pooled vaccine efficacy results reported from those studies.^1^ Fig 2b: Pseudovirus neutralization assay (PSVNA). Serum samples collected from AZD1222 vaccinees (n=19, Top panel) with six who had NAAT confirmed illness colored in light gray; and placebo recipients who had natural-infection induced antibody (n=6, bottom panel) assessed using PSVNA against the original SARS-CoV-2 D614G lineage (left), an RBD-only chimeric virus containing the K417N, E484K, and N501Y substitutions (middle) and the B1.351 lineage virus (right). Colored background indicates dilutional titre, where titers greater than 1:400 are colored dark blue, 25-400 in light blue, and titers <1:50 are colored red. Pie charts above each line graph summarize the proportion of viruses by dilutional titer. Fig 2c: Live virus neutralization assay (LVNA). Neutralization by plasma of vaccinated participants (n=19, top panel, including with NAAT confirmed illness (n=6)) and placebo recipients who had natural-infection induced antibody (n=6, bottom panel) of B.1.1 and B.1.351 variants. Participants were as for the PSVNA assay. Points are reciprocal 50% inhibitory dilution (1/ID50) per participant. Participants with NAAT confirmed illness are shaded in grey and participants with no detectable neutralization (defined as 1/ID50<1) shaded in red.

**Figure 3:**
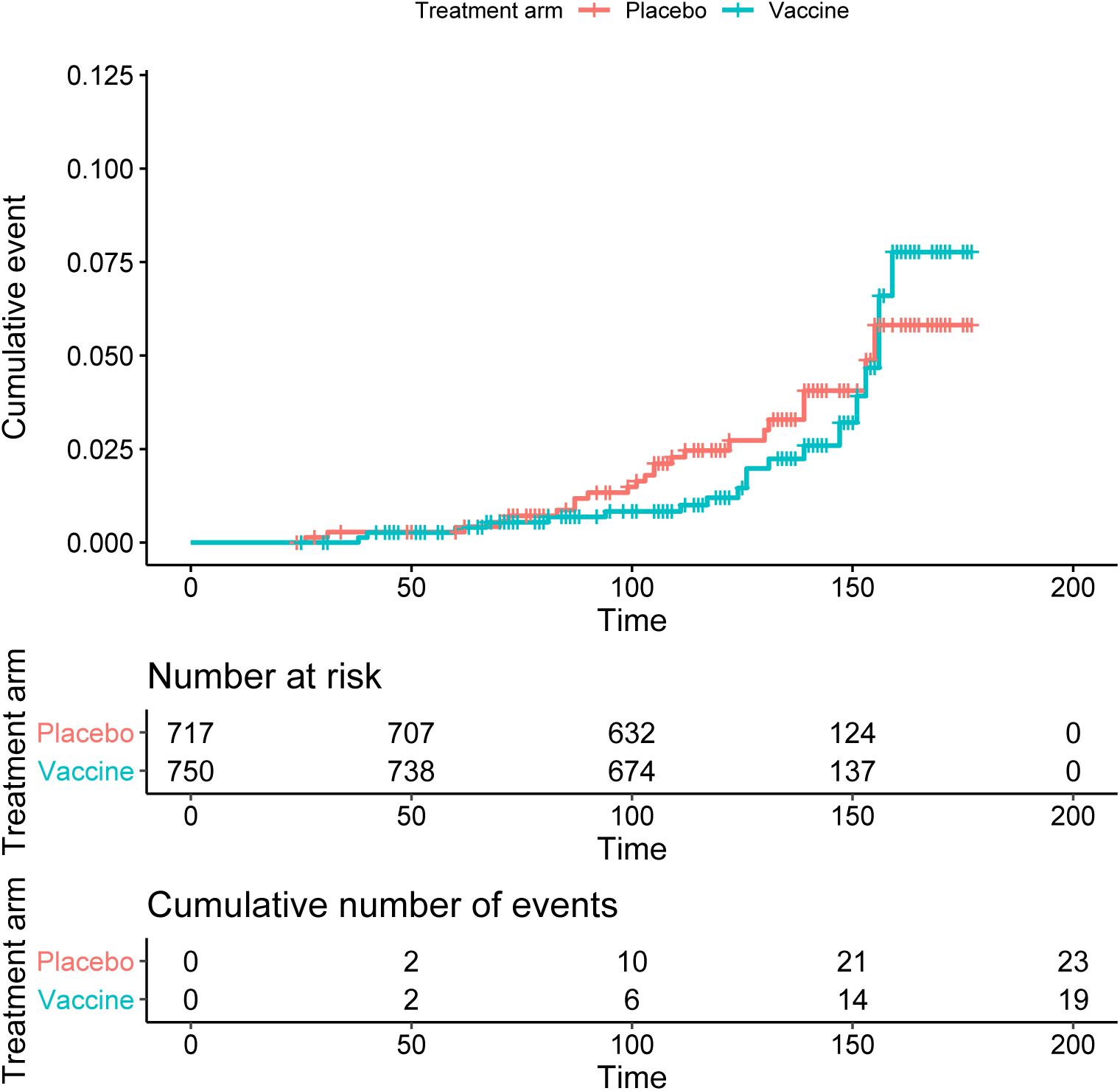
Kaplan-Meyer plot of ChAdOx-1 nCoV19 against all-severity symptomatic Covid-19 illness following two doses versus placebo.

On PSVNA testing, 9 (47%) out of the 19 seronegative vaccinees showed no neutralization activity against an RBD triple-mutant pseudovirus (containing K417N, E484K and N501Y), and 15/19 (79%) had no neutralization activity against B.1.351 pseudovirus (Fig. 2b). Vaccinees with NAAT-confirmed illness (prior to emergence of B.1.351) showed similar results to those with no NAAT confirmed illness (Fig. 2b). Samples from the SARS-CoV-2 infected placebo recipients showed similar reduction in neutralizing activity, with residual titers of <100 (or knockout) against the RBD triple-mutant and B.1.351 (Fig. 2b).

LVNA showed lower neutralization overall relative to PSVNA (Fig. 2c). Of 19 seronegative vaccinees, one had undetectable neutralization response to B.1.1 and B.1.351. Of the 18 participants with neutralization activity against B.1.1, 10 (56%) had undetectable neutralization activity against the B.1.351 variant, and the remaining eight showed a 2.5 to 31.5-fold relative reduction in neutralization (Fig. 2c). As with PSVNA, six vaccinees with NAAT-confirmed illness prior to emergence of B.1.351 showed similar results to those with no NAAT confirmed illness (Fig. 2c, light grey points). Of the 10 vaccinees with complete resistance in the LVNA, all showed complete resistance in the PSVNA. Among the six recently SARS-CoV-2 infected placebo-recipients, all demonstrated detectable neutralization of the B.1.1 variant, whereas neutralization activity against the B.1.351 variant was completely knocked out in two cases, there was 6.0-to-9.5 fold reduction in three cases and no change in one case (Fig. 2c).

Given the potential importance of T cells in protection from severe disease,^25^ we include unpublished data on a subset of 17 ChAdOx1-nCoV-19 recipients from the UK who were evaluated using TCRB sequencing for expansion of Spike specific T cells; detailed in Supplementary Text 1.8. ChAdOx1-nCoV19 resulted in expansion of both CD4^+^ and CD8^+^ specific antigens. Of 87 spike specific antigens identified by T-cell receptor variable beta chain sequencing, 75 remain unaffected by B.1.351 mutations. Of note, the D215G mutation found within B.1.351 is within a region which had prevalent T cell antigen responses (Supplementary Figure 7).

### Vaccine efficacy

All forty-two endpoint cases were graded either as mild (vaccinees=15; placebo-recipients=17) or moderate (vaccinees=4; placebo-recipients=6) with no cases of severe disease or hospitalisation in either arm. The incidence (per 1000 person-years) of Covid-19 more than 14 days after the 2nd dose among seronegative participants, and subsequent NAAT confirmed infection through to 14 days post second injection, was 93.6 and 73.1 in placebo and vaccine recipients, respectively; VE: 21.9% (95%CI: −49.9; 59.8); Table 2. Similarly, inclusive of participants that were seropositive but had a non-reactive PCR before or at randomization, the incidence (per 1000 person-years) of Covid-19 >14 days post 2^nd^ injection did not differ between placebo (81.9) and vaccine (73.2) recipients; VE: 10.6%; 95%CI:-66.4 to 52.2); Supplementary Table S5.

**Table 2:**
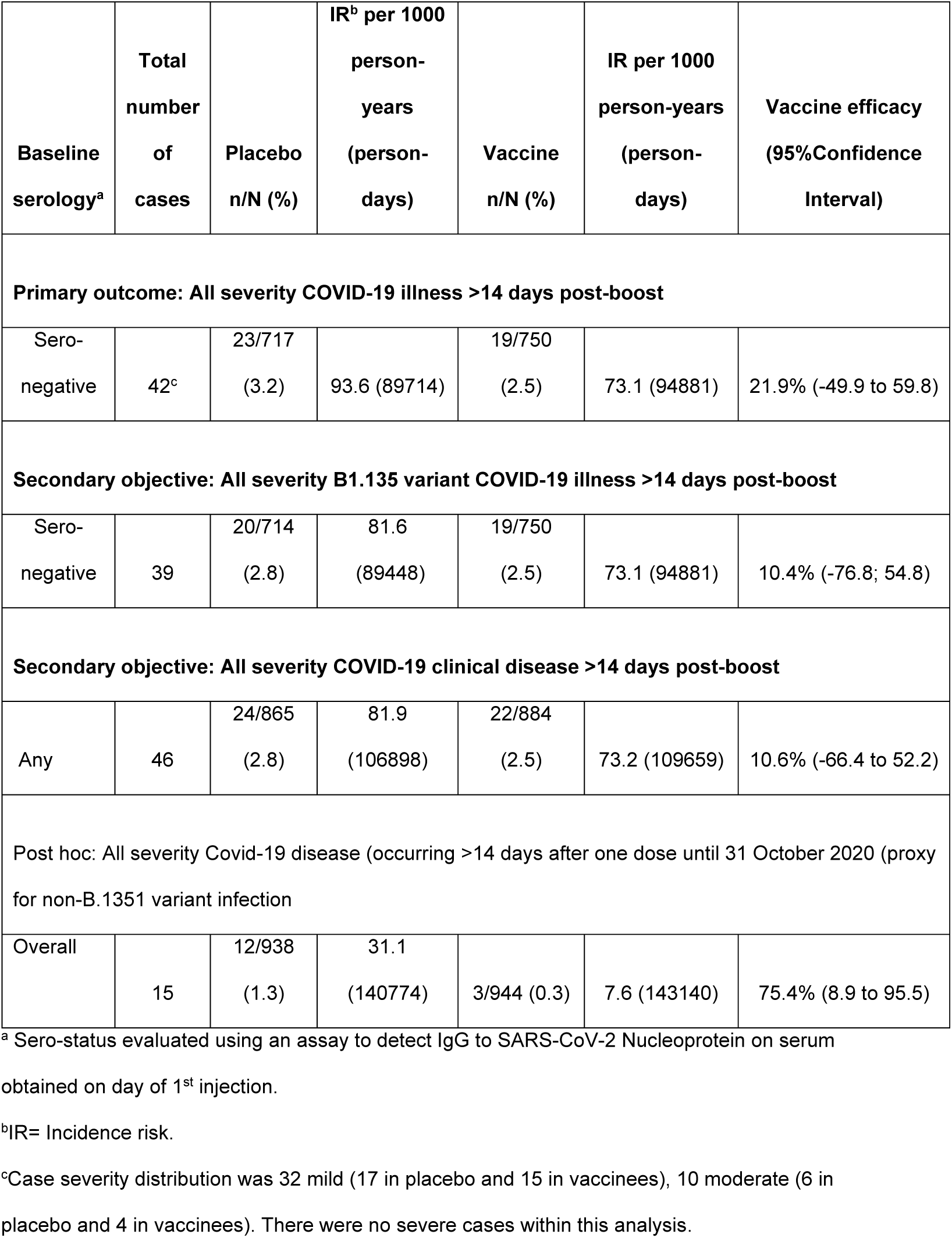
Vaccine efficacy against symptomatic Covid-19

Forty-one (97.6%) of the 42 samples of the primary endpoint cases were successfully sequenced and classified, of which 39 (95.1%) were B.1.351 and 4.9% (n=2; all in the placebo arm) were the B.1.1.1 and B.1.144 lineages; Supplementary Figure S6. Further detail of phylogenetic characterization is detailed n Supplementary text 1.7. In a secondary outcome analysis, efficacy against B.1.351 was also not evident (VE:10.4%; 95%CI: −76.8 to 54.8); Table 2.

Detailed analyses of other secondary and exploratory VE estimates are detailed in Supplementary Table S5. Overall VE for all severity Covid-19 at >14 days post-primary dose was 33.5%; 95%CI: −13.4 to 61.7. Also presented in Supplementary Table S5 are VE for any symptomatic or asymptomatic illness SARS-CoV-2 infection post-primary and post-booster injections, with VE estimates being non-significant and similar to corresponding Covid-19 disease endpoint VE.

In a post-hoc analysis, VE at least 14 days after a single dose of injection with outcomes cases limited until 31^st^ October 2020, as a proxy for infection by non-B.1.351 variant (Supplementary Figure S1)^15,26,^ the overall attack rate of mild-moderate (no sever cases) Covid-19 at least 14 days after the first injection was 1.3% and 0.3% in placebo and vaccine recipients, respectively; VE 75.4% (95%CI: 8.7; 95.5); Supplementary Table S6. Similar VE point estimates were observed for other post-hoc VE analyses for endpoints occurring until 31^st^ October 2020.

## DISCUSSION

In this study, we find 2 doses of ChAdOx1-nCoV had no efficacy against non-hospitalized mild to moderate Covid-19 mainly due to the B.1.351 variant. The lack of efficacy being specific to the B.1.351 variant, is supported by the 75% efficacy (95%CI: 8.7; 95.5) observed in our study for Covid-19 occurring at least 14 days after even a single dose of ChAdOx1-nCov19 prior to the B.1.351 variant circulating in South Africa.

Furthermore, the PSVNA and LVNA experiments provide additional evidence of reduced or abrogated vaccine-induced antibody neutralization against the B.1.351 variant. Although the degree of attenuation which compromises an effective neutralizing antibody response is unknown, the highest degree of neutralization achieved in a vaccinated participant against B.1.351 using the LVNA was a 1:20 dilution, and highest remaining titer against B.1.351 was <1:200 on the PSVNA. Comparison of the RBD triple mutant (K417N, E484K, N501Y) and B.1.351 in the PSVNA assay suggests that much, though not all, of the vaccine-elicited neutralization is directed to RBD. Similar loss of neutralizing activity of antibody induced following natural infection after the first wave of the Covid-19 outbreak against the B.1.351 variant have been recently reported.^14,17^

The extent that effectiveness of other Covid-19 vaccines may be affected by variants with B.1.351 (and P.1) like variants, could depend on the magnitude of neutralizing antibody induced by vaccination. The immunogenicity evaluation of different Covid-19 vaccines has not used a standardized assay or benchmarked against the available WHO International standard. Nevertheless, differences in fold-change between antibody concentrations post 1^st^-dose and subsequent further fold-increase following a booster dose does suggest heterogeneity in vaccine immunogenicity^.5-7,9^ The mRNA Covid-19 vaccines, although having modest neutralizing antibody after the first dose, have higher fold increase after the second dose than ChAdOx1-nCoV19 and heterologous Sputnik V (adenovirus-26 followed by adenovirus-5 vector Covid-19 vaccines)^5,6^,9. For the Moderna and Pfizer mRNA vaccines relative to activity against the D614G variant, 8.6 and 6.5 fold respective reduction in neutralization on PSVNA were observed against the B.1.351 variant, whilst no difference was evident against the N510Y.V1 (B.1.1.7)-like mutant.^19,27^

Recent interim analysis of the Novavax nano-particle spike protein Covid-19 vaccine also indicate that there may be reduced effectiveness of other Covid-19 vaccines against the B.1.351 variant.^12^ The Novavax study in South Africa included analyses for a similar spectrum of illness severity as in our study. Interim results of the Novavax Covid-19 vaccine in HIV-uninfected individuals in South Africa and the general UK population reported VE of 60% (95%CI: 20; 80; in) and 89% (95%CI:75; 95), respectively against all-severity Covid-19 (also mainly mild and moderate illness). More than 90% of the sequenced samples for primary endpoint cases in South Africa were due to the B.1.351 variant, and 57% (32/56) of the UK cases were due to the B.1.1.7 (without the E484 mutation)^12^; suggesting differences in efficacy to B.1.351 and variants without the B.1351 RBD and NTD mutations. In the absence of an established correlates of protection against Covid-19 due to the prototype or B.1.351-like variants, clinical evidence is warranted on the effectiveness of other Covid-19 vaccines against mild-moderate Covid-19 illness.

Another recent multi-national study inclusive of South Africa, evaluated efficacy of a single dose of the Janssen non-replicating adenovirus type 26 vaccine (Ad26-vaccine). Interim results from the South African arm reported VE of 54% against moderate-severe and 85% against severe Covid-19 mainly due to B.1.351 variant.^13^ The Ad26-vaccine study, however, only submitted NAAT confirmed cases that had at least three symptoms for endpoint adjudication^28^ and consequently the VE analyses likely excludes the majority of mild Covid-19 in the study. Notably, the immunogenicity of Ad26-vaccine is similar to that of ChAdOx1-nCoV19 following the first dose and after the second dose.^13,29^ The neutralizing antibody response induced by the Ad26-vaccine against the B.1.351 variant has not yet been reported on.

While there is high correlation between antibody response and VE, suggesting that the neutralizing antibody response is important, it has been reported that T cell responses may contribute to protection from COVID-19 pathophysiology even in the presence of suboptimal neutralizing antibody titers.^30^ In a post hoc inclusion in this manuscript, we report on T-helper immune responses induced by ChAdOx1-nCoV19 among vaccine recipients from the UK. In the Spike-Specific T cells that expanded following ChAdOx1-nCoV19 vaccination, the majority of antigens and epitopes remained intact to the B.1.351 variant. Hence, we speculate that extrapolating from comparable immunogenicity of ChAdOx1-nCoV19 and Ad26-vaccine, the high efficacy (85%) of the Ad26-vaccine against mainly B.1.351 variant severe Covid-19 may be due to vaccine induced T-helper lymphocyte cell responses that remain largely unaffected by mutations in the B.1.351 variant.

Although efforts are underway for a second generation of Covid-19 vaccines targeted against B.1.351 and P1 like variants, the only Covid-19 vaccines likely to be available for most of 2021 have been formulated against the prototype SARS-CoV-2. ChAdOx1-nCoV19 is likely to be one of the most accessible of all currently authorized Covid-19 vaccines^,31,32^ with expected manufacture of ∼3 billion doses during 2021 and being the least costly.^33^ Escape from human neutralizing antibody responses is expected to be a feature of the pandemic coronavirus in the years ahead as a result of pressure on the virus to select for variants that can still transmit despite immunity from natural infection or vaccination and cause mild infection. For this reason, the results reported here are expected. Nevertheless, considering the similar technology used to produce ChAdOx1 nCoV-19 and Ad26-vaccine, their comparable immunogenicity, and the high efficacy of Ad26 against severe Covid-19 mainly by B.1.351 variant in South Africa, speculatively ChAdOx1-nCoV19 may still protect against severe Covid-19. Deliberations on the utility of ChAdOx1 nCoV-19 also needs to be considered in the context of ongoing global spread and community transmission of the B.1.351 variant,^34^ and evolution of other SARS-CoV-2 lineages that include similar mutations.

## Data Availability

Data will be made available by the study statistican after acceptance of the mansucript by a journal

https://www.wits.ac.za/covid19vaccine/oxford-covid-19-vaccine-trial/.

## Contributors

Author contributions

SAM & AJP conceived the trial, SAM is the national principal investigator. SAM, AI, CLC, GK, VB and AJP contributed to the protocol and design of the study. SAM, AI, CLC, GK, VB, ALK, CT were responsible for the design and conduct of the trial, database design and development, site selection and training, data collection, data cleaning and interpretation of results.

AI, MV conducted the statistical analysis. SLB, QEB, CB, KD, LF, ALK and SDP are study site principal investigators and enrolled participants, collected data and contributed to manuscript preparation.

KA, SB, AEB, AE, MG, CH, EH, AJ, MM, MaM, MdM, KM, SO, FP, LR, CT, AT, SvE contributed to the implementation of the study at sites and/or data collection.

JNB, SH, SP, HR, HT, CKW, JG, TH, PK, LM, TM, YN and BO contributed to data generation and analysis. ML, JdP, SK, AM, SM, ML, JdP, NMD, EJK contributed to sample processing, data generation and analysis.

SAM, AI, CLC, VB, GK, PLM, AS, TdO, TLV, AP contributed to the preparation of the report. All authors critically reviewed and approved the final version of the manuscript.

## Acknowledgements

The authors would like to thanks all the volunteers who participated in this study.

The authors gratefully acknowledge the local safety physician, Guy Richards, for reviewing all SAEs.

The authors would like to acknowledge the Data safety and monitoring committee for the trial: Robert Heyderman (co-chair), Manish Sadarangani (co-chair), Paul Kaye, Steve Black, George Bouliotis, Gregory Hussey, Bernhards Ogutu, Walter Orenstein, Sonia Ramos, Cornelia L. Dekker, Elizabeth Bukusi

The authors are grateful to the independent case evaluation committee:

Jeremy Carr, Steve Chambers, Kim Davis, Simon Drysdale, Charles Feldman, Malick Gibani, Elizabeth Hammershaimb, Michael Harrington, Celina Jin, Seilesh Kadambari, Rama Kandasamy, Carla Leisegang, Toby Maher, Jamilah Meghji, Marc Mendelson, Colin Menezes, Claire Munro, Jeremy Nel, David Pace, Rekha Rapaka, Robindra Basu Roy, Daniel Silman, Gemma Sinclair, Merika Tsitsi, Jing Wang

The authors would like to acknowledge the following key trial team staff members for their valued contributions towards the trial:

Wits-VIDA: Joyce Sibuya, Rose Khoza, Nomsa Mlaba, Phindile Khumalo, Sibongile Jauza, Aletta Matywabe, Christinah Klaas, Farisai Kuonza.

Wits RHI: Gabriella Bernade, Mrinmayee Dhar, Alden Geldenhuys, Nakile Mabaso, Nomusa Msweli, Sanele Nkosi, Charmain Norman, Jean Le Roux, Tiffany Seef, Othusitse Segalo, Sarah Jane Whitaker

NICD: Bronwen Lambson, Mashudu Madzivhandila, Donald Mhlanga, Zanele Molaudzi, Frances Ayres

This report is independent research funded by the Bill & Melinda Gates Foundation (INV-017710 to SAM; INV-018944 to AS) and the South African Medical Research Council (96167).

Personnel from the KD lab were supported by the South African MRC/UCT Centre for the Study of Antimicrobial Resistance (RFA-EMU-02-2017) and the EDCTP (TMA-2015SF-1043 & TMA- 1051-TESAII). AE is supported EDCTP (TMA-2015CDF-1052)

We acknowledge funding from the South African Medical Research Council SHIP programme. P.L.M. is supported by the South African Research Chairs Initiative of the Department of Science and Technology and the NRF (Grant No 98341). C.K.W. is supported by Fogarty International Center of the National Institutes of Health under Award Number R21TW011454 and the FLAIR Fellowship program under award number FLR\R1\201782.

The views expressed in this publication are those of the author(s) and not necessarily those of the BMGF or the SAMRC

The authors are grateful to the teams at Advent (Pomezia, Italy) and COBRA Biologicals (Keele, UK) for supply of vaccines.

